# A telerehabilitation program to improve visual perception in children and adolescents with hemianopia consecutive to a brain tumour: a single-arm feasibility and proof-of-concept trial

**DOI:** 10.1101/2024.01.25.24301666

**Authors:** Mariana Misawa, Inci Yaman Bajin, Bill Zhang, Monica Daibert-Nido, Danielle Tchao, Eduardo Garcia-Giler, Kyle Cheung, Lora Appel, Pi Nasir, Arun Reginald, Uri Tabori, Ute Bartels, Vijay Ramaswamy, Samuel N. Markowitz, Eric Bouffet, Michael Reber

## Abstract

**Background:** Brain tumour in children can induce hemianopia, a loss of conscious vision, profoundly impacting their development and future prospects, yet no effective intervention exists for this pediatric population. This study aims to explore the feasibility, safety, and potential effectiveness of a home-based audiovisual stimulation in immersive virtual-reality (3D-MOT-IVR) to restore visual perception.

**Method:** In a phase 2a, open-labeled, nonrandomized, single arm study, 10 children and adolescents with stable hemianopia were enrolled to perform 20-minute sessions of 3D-MOT-IVR every other day for six weeks from home. We assessed feasibility by monitoring completion rates, remote data transfer, qualitative feedback. Safety was evaluated using validated cybersickness questionnaires. Comprehensive vision assessments were conducted pre- and post-intervention, with follow-ups at 1- and 6-month intervals.

**Results:** The home-based 3D-MOT-IVR intervention proved both feasible and safe, with no reported adverse events. All participants completed the prescribed stimulations and the pre- and post- intervention assessment points, 90% completed the follow-ups. Remarkably, the outcomes revealed significant improvements post-intervention: 50% of participants showed enhanced visual perception in their blind field, while 70% exhibited increased reading speed. Importantly, these positive effects were sustained at the 6-month follow-up. A robust correlation emerged between 3D-MOT-IVR performance and improved visual perception in the blind field, emphasizing the intervention’s effectiveness.

**Conclusion:** Our findings underscore the feasibility and safety of home-based 3D-MOT-IVR as a potential intervention for hemianopia in children. These promising results lay a strong foundation for a larger randomized controlled trial, offering hope for a meaningful breakthrough in visual rehabilitation for this vulnerable population.

**Key Points:**

- Absence of rehabilitation programs for children with visual field loss consecutive to brain tumour.
- Design of a home-based, personalized, 3D audiovisual stimulation in virtual-reality.
- Restoration of visual perception in the blind field after 3D-MOT-IVR.

**Importance of the Study:** Many children with a brain tumour suffer from visual field defects (hemianopia) dramatically impacting their cognitive and social growth with difficulties learning, limited mobility and thus restricted participation in physical activities and peer engagement. Later in adulthood, hemianopia affects social interactions and limits employment opportunities. Individuals with this condition present impaired visual scanning and exploration often associated with defective sound localization, deteriorating spatial detection. There is no visual rehabilitation intervention for children with hemianopia. This feasibility/proof-of-concept trial showed that a dynamic audiovisual stimulation in virtual-reality conducted every other day for 6 weeks is a feasible, safe and acceptable intervention, restoring visual perception in the blind field of 50% of the participants and improving activities of daily living. The personalized intervention was administered at home through a remotely controlled virtual-reality device, reducing the burden of disease by limiting in clinic visits and providing specialized care to children living outside urban areas.

## Introduction

A brain tumour and its treatment can affect the visual system^1,2^. In a recent study from the Children Cancer Survivor Study, 22.5% of survivors of childhood astroglial tumours ≥5 years post-diagnosis had visual impairment^3^. Visual impairment includes decreased visual acuity and contrast sensitivity, colour vision loss, and visual field loss, which are often underdiagnosed. Seventy nine percent of individuals with brain tumours who have visual field defects present hemianopia (homonymous, bitemporal or quadrantanopia)^1,2^, a loss of conscious vision caused by direct or indirect damage to the primary visual cortex V1, optic pathway, and midbrain/thalamus (through compression, infiltration or surgical intervention)^4^. The most common tumours associated with hemianopia are low-grade gliomas, craniopharyngiomas, and germ cell tumours, which otherwise have excellent prognoses with 10-year survival rates above 90%^1,2^, but most survivors are visually impaired. Although natural recovery of hemianopia occurs in 40% of the patients within 6-18 months, it is often incomplete and limited, and still leaves 60% of individuals with permanent visual field loss^2,5^. Individuals with hemianopia present long term difficulties in detecting stimuli in the defective visual field and show impaired visual scanning and exploration^6^ often associated with impaired localization of sound therefore worsening spatial detection^7,8^. From a quality of life standpoint, hemianopia affects children and adolescents in their daily living activities, including school, physical activities, and social life, profoundly impacting intellectual and social development, mobility and independence. It is also a potential source of danger (crossing streets, bumping into people or objects) and later in adulthood, can prevent these patients from driving and can affect access to employment opportunities. Altogether, there is growing recognition of the diverse and deep impact of hemianopia on physical and mental health, quality of life, and social outcomes of the affected children, the adult they become and their family. However, despite the frequent impact of brain tumours on vision, ophthalmologic evaluations are not the standard of care for all patients and there are no standardized protocols for vision loss management in the paediatric population with hemianopia^2^. Sparse evidence shows that visual acuity and visual field deficits can be improved with Bevacizumab, an inhibitor of vascular endothelial growth factor, in children with optic nerve gliomas. However, the benefits are not sustained after discontinuation of the drug and there is potential toxicity^9–12^. In our previous case series^13,14^, we used immersive virtual-reality (IVR) in the Oculus Go headset to deliver a personalized, remotely controlled, dynamic audiovisual stimulation in young adult patients with stable hemianopia (> 18 months) consecutive to a paediatric brain tumour. We demonstrated, after 6 weeks of repeated, home-based audiovisual stimulation, a clinically and statistically significant improvement in visual perception in the blind hemifield (note that 2 patients followed home-based stimulation remotely controlled from the laboratory during the 2020-2021 COVID-19 pandemic)^13,14^. These encouraging results prompted us to further investigate the feasibility, safety and potential effectiveness of our audiovisual stimulation intervention in real-world condition in the paediatric population.

## Materials and Methods

### Study Design

The study was designed as a single-arm phase II feasibility and proof-of-concept trial with pre/post-intervention analysis. The study protocol was approved by the Research Ethics Boards at the Hospital for Sick Children in Toronto, Canada (Sickkids REB# 1000076413) and at University Health Network, Toronto, Canada (UHN REB# 21-5978) and registered on clinicaltrials.gov (NCT05065268) following the principles outlined in the World Medical Association Declaration of Helsinki. All participants provided written informed consent. The study took place from July 2022 to October 2023.

### Participants (table 1)

Inclusion criteria were: patients 8-18 years old, diagnosed with homonymous hemianopia consecutive to a pediatric brain tumour, stable for > 18 months, visual acuity (BCVA) ≥ 20/200, interpupillary distance ≥ 56mm, ability to follow the visual and auditory stimuli and training instructions, home Wi-fi access. Concomitant treatment with chemotherapy or targeted therapy was allowed with the following conditions: on treatment for > 6 months and no change in visual assessment in the last 6 months.

**Table 1:** Participants information and demographics. HH: Homonymous Hemianopia, BTH: Bitemporal Hemianopia.

| Participant | Sex at Birth | Age range (years) | Time From Diagnosis (months) | Visual Field deficit | Tumour type | Location of Tumour | Active Chemo/target Treatment |
| --- | --- | --- | --- | --- | --- | --- | --- |
| P1 | F | 15-19 | 108 | Right HH + BTH | Ganglioglioma | Chiasmatic | Yes |
| P2 | M | 15-19 | 120 | Right HH | Pilocytic astrocytoma | Chiasmatic | Yes |
| P3 | F | 15-19 | 156 | Right HH | Pilocytic astrocytoma | Posterior | Yes |
| P4 | M | 15-19 | 72 | Left HH | Pilocytic astrocytoma | Posterior | No |
| P5 | M | 15-19 | 192 | Left HH | Pilocytic astrocytoma | Chiasmatic | No |
| P6 | M | 10-14 | 48 | Right HH | Glioblastoma-IDH wildtype | Posterior | No |
| P7 | M | 10-14 | 72 | Right HH | Pilocytic astrocytoma | Chiasmatic | Yes |
| P8 | M | 10-14 | 96 | Right HH | Diffuse astrocytoma, MYBL1-Altered | Posterior | No |
| P9 | M | 15-19 | 24 | Left HH | Pilocytic astrocytoma | Chiasmatic | No |
| P10 | F | 10-14 | 72 | Left HH | Supratentorial YAP1 Fused ependymoma | Posterior | No |

Exclusion criteria were: pre-existing or ongoing ocular disease, both eyes with media opacity that impairs microperimetry testing, inability to perform during testing and training, consumption of psychoactive drugs, 3 consecutive virtual-reality symptoms and effect (VRISE) questionnaire scores < 25 at inclusion indicative for cybersickness^15^, history of vertigo or dizziness, prior visual rehabilitation interventions.

### Intervention plan: Dynamic audiovisual stimulation procedure

The participants followed a dynamic audiovisual stimulation procedure in IVR at home using the standalone and remotely controlled headset Meta Quest 2/Pro for 4 (P1-P2) or 6 weeks (P3-P10), corresponding to 13 or 21 sessions for a total time of 3 h 15 min or 5 h15 min of stimulation, respectively. Every other day, participants performed 1 session of stimulation composed of 3 blocks of 15 trials of 20 s, equivalent to 3 x 5 min of continuous audiovisual stimulation. The audiovisual stimulation task corresponds to the multiple object-tracking (MOT) paradigm, developed in the 80’s to study visual attention in humans^16,17^. We implemented a 3D version of the MOT in virtual-reality, encoded in the Meta Quest 2/Pro using 3D Unity programming platform. The 3D-MOT is composed of 8 high-contrast orange spheres on a black background (luminosity = 100 cd/m^2^, size = 1.57° of visual angle). After one random sphere was cued (turning blue for 5 seconds and switching back to orange, Figure 1), the spheres move for 20 seconds following random linear paths, bouncing on one another and on the walls of a virtual 3D cube when collisions occur (Figure 1). The overall span of the movement of the spheres covers 80° and 70° of horizontal and vertical visual angle respectively as measured using augmented reality and a geometric chart. The initial speed of the spheres was adjustable, from 3 °/s to 240 °/s and determined individually for each participant at baseline. Spatial sound was also emitted by the cued target and was spatially and temporally correlated to its movement. The sound encoded corresponded to a continuous (a loop function in the audio source setup), low pitched sound (60 Hz, 50 dB Hearing Level). The maximum delay between sound and target was equivalent to the refreshing rate of the headsets corresponding to 16.6ms (temporal tolerance), below the 100ms temporal disparity above which multisensory response enhancement decreases^18^. After 20 s, the movement of the spheres stopped and the participants were asked to select, using a virtual laser pointer, the cued sphere (the target) among the other spheres (mark-all procedure)^17^. If the selection was correct (corresponding to the cued target) a positive feedback sound was provided and the speed of the spheres in the next trial increased. If the selection was incorrect (corresponding to a distractor) a negative feedback sound was provided and the speed of the spheres in the next trial decreased. Speed was modified following an adaptative simple up-down staircase method: speed variation X: X_n_ = X_(n-1)_ +/- [X_(n-1)_/n]^19,20^. During the first 2 weeks of the intervention (7 sessions) participants were allowed to use head/eye movements to track the moving target (overt tracking). In the following 2 or 4 weeks (7 or 13 sessions), participants were instructed to keep their gaze still (head and eye fixated on a fixation point in the center of the display) to track the moving target with their peripheral vision (covert tracking). To control for fixation and to investigate tracking strategy during the intervention, head/eye position was recorded during the task by the Meta Quest 2/Pro head/eye trackers. Head tracking was downsampled at 10 Hz. Eye tracking was sampled at 90 Hz (accuracy = 1.65 °, precision = 0.7 ° in head-free condition)^21^ after 9-point calibration.

**Figure 1:**
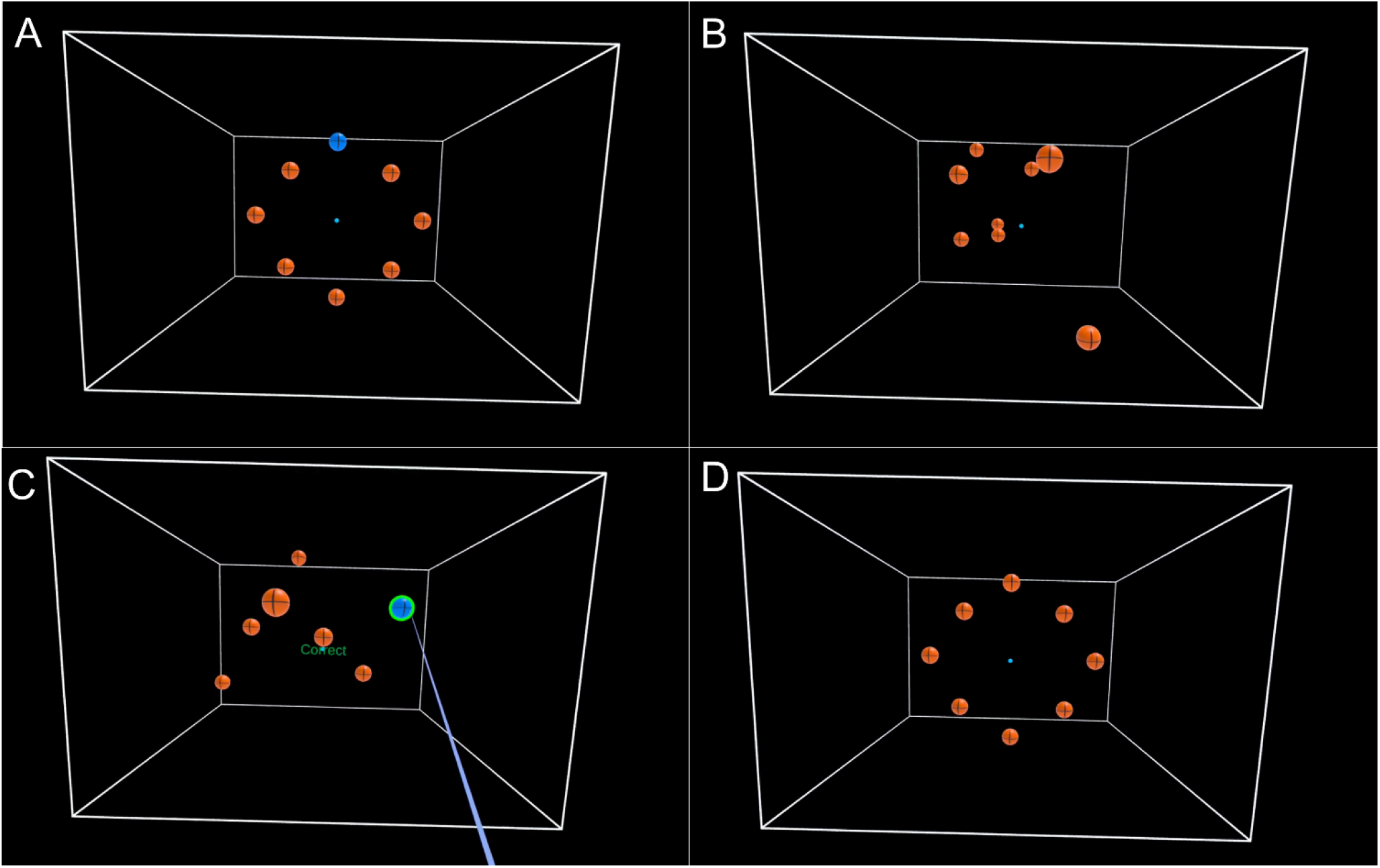
Dynamic audiovisual 3D-MOT-IVR stimulation. **A**. 8 orange spheres appear in the headset. One sphere is arbitrary cued (blue) and returns to orange after 5 seconds. **B**. All spheres start moving randomly for 20 seconds. **C**. After 20 seconds, the spheres stop moving and the participants must select, using laser pointer (light blue) the sphere matching the one previously cued (mark all procedure). The outcome is then displayed as either a hit (indicating a correct selection) or a miss (reflecting an incorrect selection), accompanied by feedback sound. **D**. Sequence returns to start configuration for the next trial.

### Safety, monitoring, criteria for removal, early stopping rules, condition of replacement

The monitoring of the 3D-MOT-IVR stimulation program performed at home occurred after each block of 5 min. when the headset sent data relative to the participants’ adherence and compliance (# of task and # of block initiated and completed, duration of the blocks/session), performance (correct/incorrect selection ratio, reaction time, speed of the target, head movement) and safety (VRISE scores before and after intervention) to our secured laboratory computer through Wi-Fi. Criteria for stopping or withdrawing participants were 3 consecutive home-based VRISE scores post-intervention < 25^15^. There was no replacement for withdrawn participants.

### Outcome measures

Primary outcomes corresponded to feasibility and safety and are detailed in Table 2 and Supplementary Table 1. Secondary outcomes corresponded to pre- and post-intervention assessments of visual function and functional vision performed at the Ophthalmology Low vision Clinic at the Toronto Western Hospital (University Health Network, Toronto, Canada) following standard procedures in low-vision rehabilitation^22,23^. These assessments included visual acuity (best corrected visual acuity - BCVA) measured using the Early Treatment Diabetic Retinopathy Study (ETDRS) charts at 4 m (far vision) and the Colenbrander English continuous text near vision cards at 40 cm (near vision); contrast sensitivity (CS) measured using the Functional Acuity Contrast Test (FACT). Fixation stability (FS, Bivariate contour ellipse area, BCEA 63%) was measured using the Macular Integrity Assessment (MAIA) microperimeter (CentreVue, Padova, Italy). Fields of vision were evaluated by the monocular Humphrey full field test (Full Field 120 points Suprathreshold test) using the Humphrey Full Field Analyzer 3 standard automated perimeter (HFA 3, Zeiss, Heilberg, Germany). All these tests were performed on both left (LE) and right (RE) eye. The Esterman binocular field test (Esterman Binocular Suprathreshold test) was performed using the HFA 3 automated perimeter. Reading speed was measured at critical print size using the Minnesota Low Vision Reading (MNREAD) test. Quality of life/vision was measured using the Children’s Vision Function Questionnaire (CVFQ - 48 questions). These tests were performed binocularly.

**Table 2:** Feasibility outcome measures.

|  | Protocol completion (# of subjects) | % of IVR session completion | Safety - VR sickness questionnaire score | Dropouts |
| --- | --- | --- | --- | --- |
| Criteria | $\geq 8/10$ | $\geq 80\%$ | $\leq 3$ consecutive VR $< 25/35$ | $\leq 2$ |
| Results | 10/10 | 91.2% $\pm 6$ | - 1 VRISE score $< 25$ post-intervention.<br>- Average VRISE score post-intervention: 33.1 $\pm 2.8$. | 1 withdraw post-intervention |

Exploratory outcomes corresponded to participants’ home-based performance at the 3D-MOT-IVR task (target speed for correct choice, time of the day) and head/eye tracking strategy (head/eye movements during 3D-MOT-IVR). Performance and head tracking were measured using the Meta Quest 2 headset in all participants during each session performed at home. Eye tracking was measured using the Meta Quest Pro headset (released in Canada in November 2022) at baseline, end-of-overt tracking and at the end-of-intervention visits at the clinic for P8, P9 and P10. Tracking strategy was investigated using correlation of head/eye trajectory with target trajectory calculated after vectorization of target, head and eye movements (coordinates related to time along the horizontal and vertical axes).

### Data Analysis

Monocular BCVA was converted into the logarithm of minimal angle of resolution scale (LogMAR) for analysis purposes. Eye/head positions as a function of time were recorded in a .csv file sent through Wi-fi to our lab computers. Data extraction, analysis and representation was performed using in-house codes in Python. Head/target and head/eye/target trajectory correlation were calculated after vectorization (Δcoordinates/Δtime) using Python Person’s correlation function. Data related to the CVFQ were collected but not processed because the parents must be the responders of the CVFQ^24^. However, in the majority of cases, we could not control for questionnaires responders and questionnaires were randomly answered by either the mother or the father creating strong discrepancies in the responses. We therefore decided not to analyse this outcome measure.

### Statistical Analysis

Data analysis was based on descriptive statistics including a measure of a central tendency (median) and dispersion (min, q1, q3, max). Statistical comparisons between pre- and post-intervention data were performed using both frequentists and Bayesian paired samples t-test in JASP (v0.17.2.1) using the default effect size priors (Cauchy scale 0.707). Results were reported using the one-tailed Bayes factor BF_+0_ that represents p(data|H+:post-intervention > pre-intervention) / p(data|H0: post-intervention = pre-intervention) and two-tailed BF_10_ representing p(data|H1: post-intervention ≠ x month follow-up) / (p(data|H0: post-intervention = x month follow-up) and p(data|H1: 1 month follow-up ≠ 6 month follow-up) / (p(data|H0: 1 month follow-up = 6 month follow-up). Effect sizes estimates were reported as median posterior Cohen’s δ with 95% credibility interval. Linear mixed model (LMM) analysis was used to analyse random effects on the Esterman binocular field test and on the performance at 3D-MOT-IVR. Visual assessment results were analyzed using the coefficient of repeatability (COR), specific to each assessment, comparing pre- and post-intervention data^25^. The COR (or smallest real difference) quantifies an absolute reliability in the same unit as the assessment tool and is directly related to the 95% limits of agreement proposed by Bland and Altman^26^. It corresponds to the value below which the absolute differences between two measurements would lie within 95% of probability. Therefore, measurement values strictly different from COR indicate an effect of the intervention^25,27^.

## Results

### Feasibility - Safety

Feasibility criteria were defined based on our previous case series^13,14^, studies using virtual-reality as an interventional tool^28,29^ and CONSORT guidelines for feasibility trials^30^ (Table 2). Results indicated that 100% (10/10, protocol completion criteria ≥ 80%, Table 2) of the participants performed 91.2% (session completion criteria ≥ 80%, Table 2) of the home-based scheduled sessions (P1-P2 13.5 sessions/15, P3-P10 19.1 sessions/21). Safety, assessed using the VRISE questionnaire scores evaluating cybersickness, showed an average of 33.1 out of 35 points (and 1 score < 25 points) indicating no adverse event (cybersickness symptoms scores criteria ≤ 3 consecutive VRISE scores < 25) related to the use of the 3D-MOT-IVR^15,31^ (Table 2). Detailed analysis showed that among the 5 types of VRISE symptoms (nausea, disorientation, dizziness, fatigue and instability scored from 1: severe to 7: no symptoms), fatigue was the most pronounced symptom after intervention (summed difference score ΣΔ = - 84) followed by dizziness (ΣΔ = - 45), nausea (ΣΔ = - 36), disorientation (ΣΔ = - 32) and instability (ΣΔ = - 17) for a total of n = 180 sessions. No drop-outs due to the intervention were recorded although 1 voluntary withdraw was recorded due to time constraints of visual assessments at the clinic at 1- and 6-month follow-up.

Feasibility of remote data collection indicated that 100% (180/180) of the sessions performed at home by the participants were recorded on the headset and that 100% (180/180) of the corresponding reports (target speed for correct choice, time of the day, head tracking and VRISE scores) were transferred through Wi-fi and received on the lab computers, indicating no loss of digital data. Qualitative feedback was performed at the end of the intervention period using an adapted version of the virtual-reality neuroscience questionnaire (VRNQ)^15^. Median score (43 [37, 45, 50, 53]) indicated usability of the 3D-MOT-IVR stimulation (total cutoff threshold = 40, total max score = 56, Supplementary Table 1). Detailed analysis indicated that, among 8 items (scored 1: extremely difficult/low to 7: extremely easy/high, cutoff threshold = 5)^15^, quality of the sound (5 [3, 5, 5.75, 7]), quality of the graphics (5 [4, 4.25, 5, 6]) and usability of the laser pointer (5 [4, 4.25, 7, 7]) had the lowest scores, although above cutoff threshold (Supplementary Table 1).

### Preliminary effect of the intervention

Esterman binocular field testing was assessed pre- and post-intervention. Results showed significant evidence for a increase of the number of points perceived (Δ = + 3.4 points ±1.4, COR = ±3^14^) in the blind field in 50% of the participants (5/10, responders) after intervention (pre-intervention: 72.0 [60.0, 71.0, 75.0, 114.0], post-intervention: 78.0 [61.0, 73.0, 80.0, 117.0], t_(4)_ = 2.81, p = 0.048, BF_+0_ = 4.38, δ = 0.91, [95%CI, 0.10, 2.17], Figure 2A, B). Non-responders (5/10) showed no changes between pre- and post-intervention (Supplementary Figure 1A). Follow-up analyses revealed no difference between post-intervention and 1- and 6-month follow-up in both responders and non-responders (Figure 2A, Supplementary Figure 1A). Fixation monitoring by the healthcare provider during the Esterman binocular field test revealed no loss of fixation above threshold (20%). All responders (5/10) followed a 6-week intervention whereas 2 participants (P1 and P2) followed a 4-week intervention and did not show visual field recovery. This indicates that 5/8 participants following a 6-week intervention showed visual field improvement.

**Figure 2:**
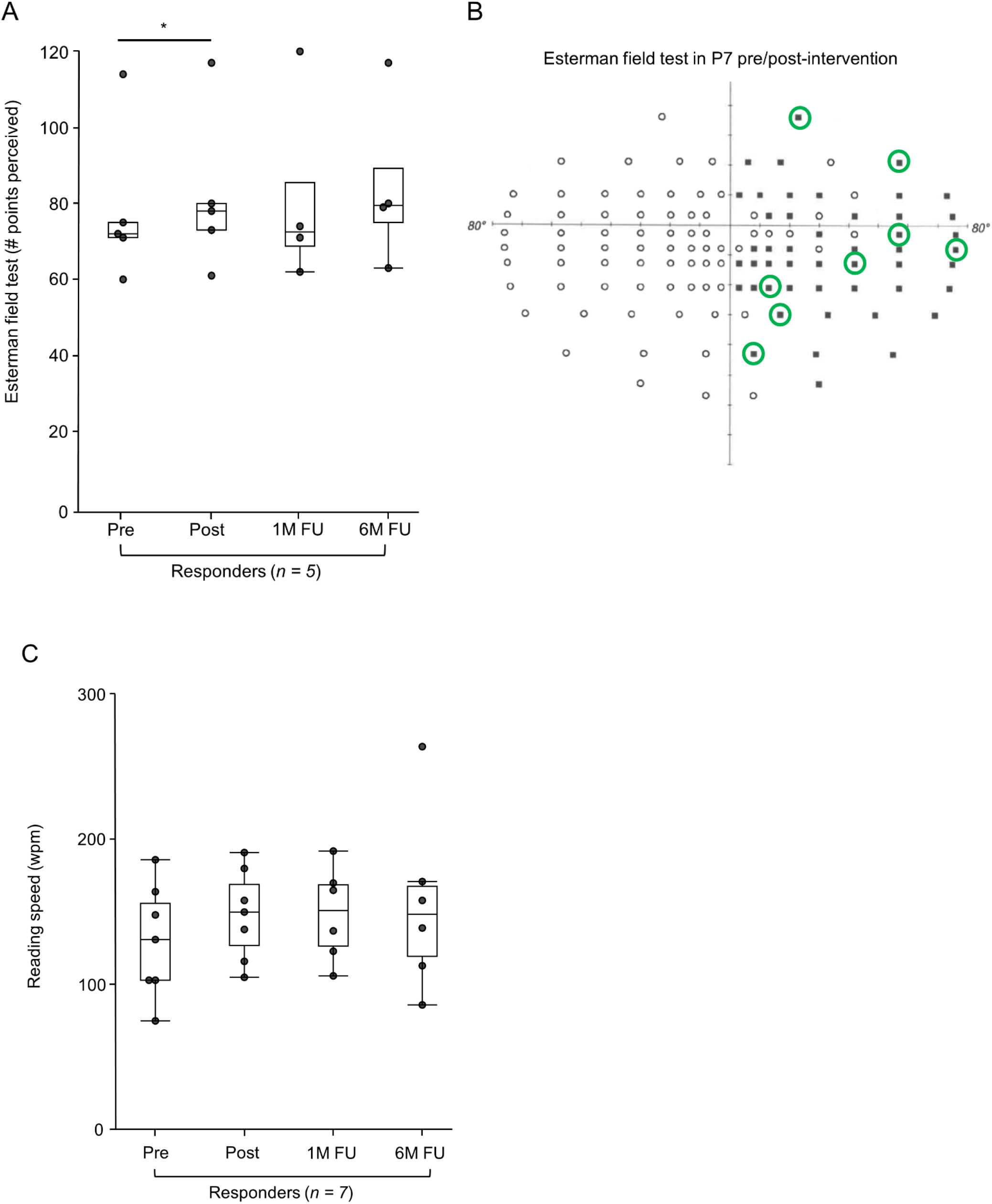
Secondary outcomes. **A.** Number of points seen at the Esterman binocular field test in responders (n = 5/10) pre/post-intervention and at 1-month (1M FU) and 6-month (6M FU) follow-up (1M FU: 72.5 [62.0, 68.7, 85.5, 120.0], 6M FU: 79.5 [63.0, 75.0, 89.2, 117.0], post x 1M FU: t_(3)_ = 0.322, p = 0.769, BF_+0_ = 0.542, δ = 0.30, [95%CI, 0.01, 1.03]; post x 6M FU: t_(3)_ = 1.608, p = 0.206, BF_+0_ = 0.232, δ = 0.16, [95%CI, 0.00, 0.68]; 1M FU x 6M FU: t_(3)_ = 1.162, p = 0.329, BF_+0_ = 0.253, δ = 0.17, [95%CI, 0.00, 0.71], (*p < 0.05, BF_+0_ > 3). **B.** Representative example of a binocular Esterman field test results pre- (white circle – point perceived and black squares – points not perceived) and post-treatment (green circles) in P7. Green circles indicate the points perceived after treatment (+8 points among 48 originally not seen in the blind field: Δ = + 16.7%). **C.** Reading speed in words per minute (wpm) in responders (n = 7/10) pre/post-intervention and at 1-month (1M FU) and 6-month (6M FU) follow-up (1M FU: 151 [106, 126, 168,192], 6M FU: 148 [86,119, 168, 264], post x 1M FU: t_(5)_ = 0.115, p = 0.913, BF_+0_ = 0.34, δ = 0.29, [95%CI, 0.01, 0.91]; post x 6M FU: t_(5)_ = 0.284, p = 0.788, BF_+0_ = 0.46, δ = 0.26, [95%CI, 0.01, 0.86]; 1M FU x 6M FU: t_(5)_ = 0.462, p = 0.663, BF_+0_ = 0.53, δ = 0.29, [95%CI, 0.01, 0.91]).

Reading speed showed anecdotal evidence for a medium increase (Δ = + 18.3 ±9.6) of the number of words per minute (wpm) in 7/10 participants (responders) after intervention (pre-intervention: 131 [75, 103, 156, 186], post-intervention: 150 [116, 127, 169, 180], t_(6)_ = 1.91, p = 0.106, BF_+0_ = 2.14, δ = 0.57, [95%CI, 0.05, 1.38], Figure 2C). Although there was no significant statistical evidence, an increase of 18.3 wpm in reading speed is clinically significant (COR ±8.6 wpm)^32^. Non-responders (3/10, who are also Esterman non-responders) did not show any changes between pre- and post-intervention (Supplementary Figure 1B). Follow-up analyses revealed no difference between post-intervention and 1- and 6-month follow-up in both responders and non-responders (Figure 2C, supplementary figure 1B).

No differences were detected in far (4 m) and near (40 cm) distance Best-Corrected Visual Acuity (BCVA, LogMAR) pre/post-intervention (Supplementary Table 2A). Among the participants, two exhibited moderate visual impairment at a far distance (P2-RE = 0.7 and P5-RE = 0.6, and P5-LE = 0.8, as per the World Health Organization definitions where 0.5 < LogMAR ≥ 1). Additionally, two participants were classified as blind (LogMAR > 1.3) in one eye at 4 m (P9-RE = 1.9, P1-LE = 2.4). Notably, BCVA at 40 cm was considered normal (LogMAR < 0.5) in 8 out of 10 participants. However, two participants were identified as blind in one eye (P9-RE = 1.9, P1-LE = 2.4). No differences were noted during the 1- and 6-month follow-ups concerning far and near BCVA (Supplementary Table 2A). Fixation stability values, except for 2 participants (P1-LE = 4.72 °^2^ and P9-RE = 20.1 °^2^), were within the normal range (normal BCEA 63% = 0.8 °^2^ ±0.68)^33^ before intervention. No differences were observed pre/post-intervention at the group level (Supplementary Table 2B). Post-intervention, P1-LE and P9-RE showed clinical improvement in fixation stability (P1-LE = 1.17 °^2^ and P9-RE = 8 °^2^, COR ±0.61^14^). However, at the 1- and 6-month follow-ups, these benefits were partially lost (1-month: P1-LE = 3.8 °^2^ and P9-RE = 10.9 °^2^, 6-month: P1-LE = 2.6 °^2^ and P9-RE = 8.7 °^2^).

Contrast sensitivity (CS) values (logit) in both eyes were in the normal range pre-intervention for 7/10 participants. Statistically significant increases were observed pre/post-intervention, however, these variations were below the frequency-specific COR values and therefore not clinically significant^34^ (Supplementary Figure 2A). Three participants had impaired CS at specific frequencies (P5-RE: 1.32 (2 cyc/°), 0 (4 cyc/° and 6 cyc/°), P9-RE: 0 at all cyc/°, P1-LE: 0 at all cyc/°) which improved for P5 and P9 but not for P1 after intervention (P5-RE: 1.65 (2 cyc/°), 0.9 (4 cyc/°), 0.6 (6 cyc/°), P9-RE: 0.48 (0.5 cyc/°), 0.06 (1 cyc/°), 0.7 (2 cyc/°)). No changes were observed at 1- and 6-month follow-up.

The Humphrey monocular visual field measures were invalid and disregarded in 6/20 (30%) tests pre- intervention (3 false negative > 33%, 3 fixation losses > 20%), in 2/20 (10%) tests post-intervention (2 fixation losses > 20%), in 3/18 (16.7%) at 1-month and in 2/18 (11%) at 6-month follow-up. Using the remaining valid data (63/76 for 9 participants), the number of points perceived pre-intervention (69.5 [0, 50.7, 77.2, 91] out of 120 points) did not improve after intervention (67 [0, 61, 74.7, 84], Supplementary Figure 2B). No changes were observed at 1- and 6-month follow-up (Supplementary Figure 2B).

### Exploratory outcomes

The potential use of VR as a home-based interventional tool is innovative, therefore data about its use in real-world conditions is still lacking. We recorded the time of the day when participants were performing the 3D-MOT-IVR sessions at home and showed that the majority of the sessions (55%, n = 98) were performed between 15:00 and 21:00 (Figure 3A) taking an average time of 30 min 14 s ±4 min 28 s to complete one session (includes pre-intervention VRISE questionnaire - 3 blocks of 15 trials of 20 s each – post-intervention VRISE questionnaire). There was no effect of the time of day of the intervention on performance (Time of the day intervention x Performance: F_(5, 4.15)_ = 0.55, p = 0.738, Figure 3A).

**Figure 3:**
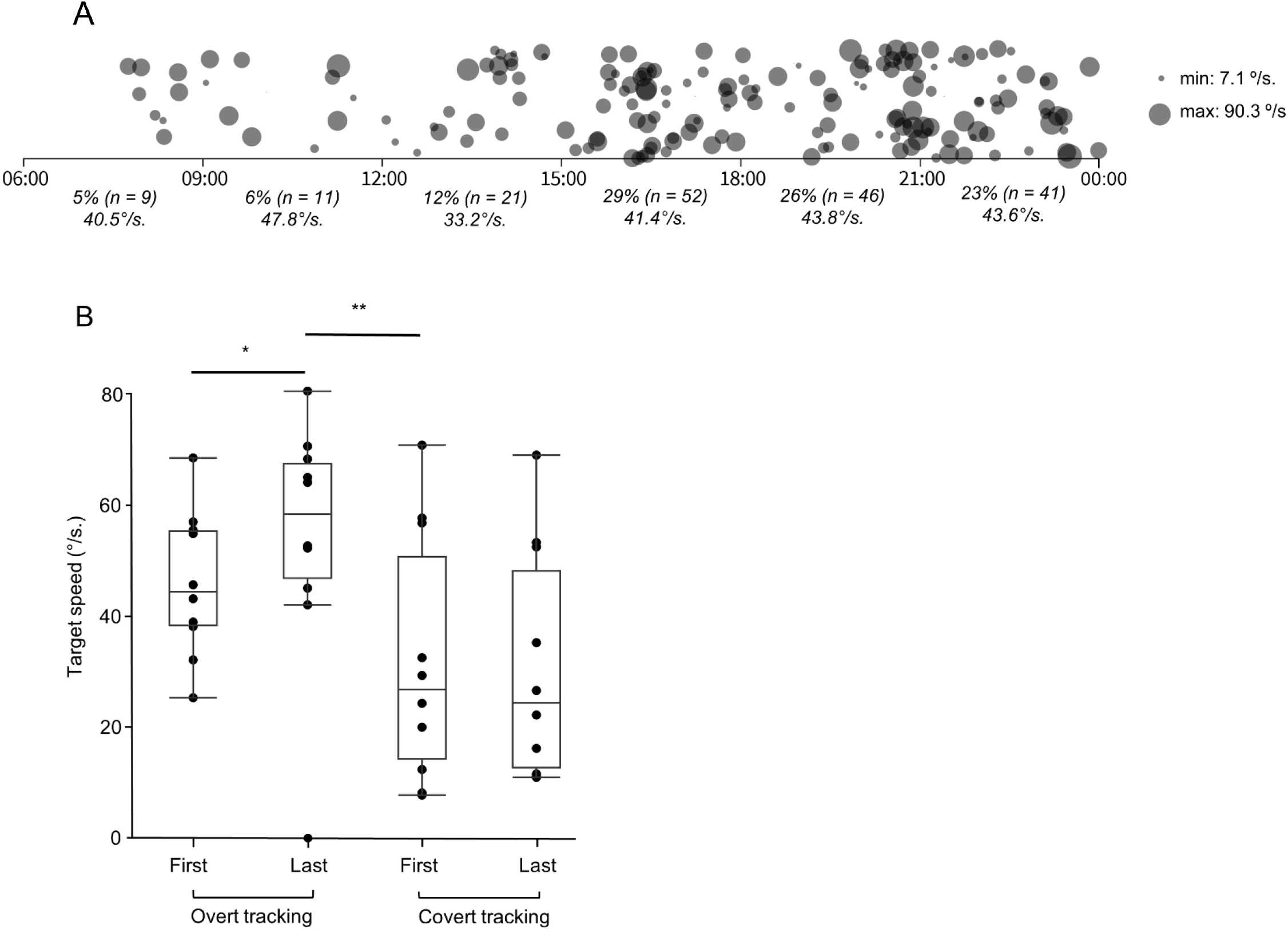
Exploratory outcomes. **A**. Distribution and number of sessions (n = 180) performed during the day (06.00 – 00.00 divided in 3 hours bins) by the participants (n = 10). The size of the circle indicates the performance of the session (average target speed in °/s for a correct choice). **B**. Graph representing the target speed for correct choice in the participants (n = 10) during the first and last session of overt and covert tracking (*: p < 0.05, 3<BF_+0_<10. **: p < 0.01, 10<BF_-0_<30).

We next explored factors that could have influenced the response of the participants (responders versus non-responders) at the Esterman field testing after the intervention. There was no effect of tumour location (posterior versus chiasmal), treatment status (active treatment versus no active treatment) or time from diagnosis (2 to 16 years) on the number of points perceived (Tumour location x Intervention x Responders: F_(1, 6.0)_ =0.03, p = 0.868; Treatment status x Intervention x Responders: F_(1, 6.0)_= 0.081, p = 0.785; Time from diagnosis x Intervention x Responders: F_(1,6.0)_ = 0.30, p = 0.606).

Participants followed the same 3D-MOT-IVR stimulation program (2 weeks of overt tracking followed by 2 weeks -P1, P2- or 4 weeks -P3 to P10- of covert tracking). We measured the performance (speed of the target for a correct choice) during both overt and covert tracking paradigms. We showed moderate evidence for medium increase (Δ = +13.1°/s ±4.7) of the target speed between the first and last session of overt tracking (when head/eyes can move to track the target) in all participants (t_(9)_ = 2.74, p = 0.028, BF_+0_ = 6.33, δ = 0.72, [95%CI, 0.12, 1.46]) as previously reported^17^ (Figure 3B). By contrast, there was no difference of target speed between the first and last session of covert tracking, (when gaze is fixed on a central fixation point and target tracked using peripheral vision, t_(9)_ = - 0.44, p = 0.669, BF_10_ = 0.336, δ = - 0.11, [95%CI, - 0.67, 0.44]) (Figure 3B). There was strong evidence for a large decrease (Δ = - 27 °/s ±7.0) in the target speed between overt and covert paradigm (t_(9)_ = −3.62, p = 0.006, BF_-0_ = 19.8, δ = - 0.96, [95%CI, - 1.80, - 0.23], Figure 3B).

To ensure participants’ compliance and explore tracking strategies, we recorded head movements every 100 ms for 20 s trials (n = 8,100 recordings) and eye movements every 14 ms for 20 s trials in 3 participants (P8-P10, n = 240 recordings). In overt tracking, eye/target correlation was high (r = 0.72 [- 0.89, −0.22, 0.90, 0.96]), while head/target correlation was lower (r = 0.25 [-0.74, −0.03, 0.51, 0.95]), suggesting reliance on eye movement with no/less contribution of head movement, consistent with previous findings on visual orienting^35^ (see example Figure 4 A, A’). Importantly, covert tracking showed low eye/target and head/target correlations (r = 0.27 [-0.81, −0.06, 0.50, 0.93] and r = 0.04 [-0.94, −0.22, 0.29, 0.89] respectively), indicating that participants maintained fixation on the central point and did not move their eyes to track the target (eye/target correlation overt versus covert: t_(239)_ = 8.62, p = 4.72E- 16, BF_+0_ = 1.14E13, δ = 0.55 [95%CI: 0.41, 0.69], example Figure 4B, B’). Notably, when correlated to participants’ blind field, results indicated that they successfully tracked the target without moving eyes or head (average correct target selection [76.8% ±9.6] above chance [1/8 = 12.5%], example Figure 4B’ – corresponding to correlation data), reinforcing the validity of our observations.

**Figure 4:**
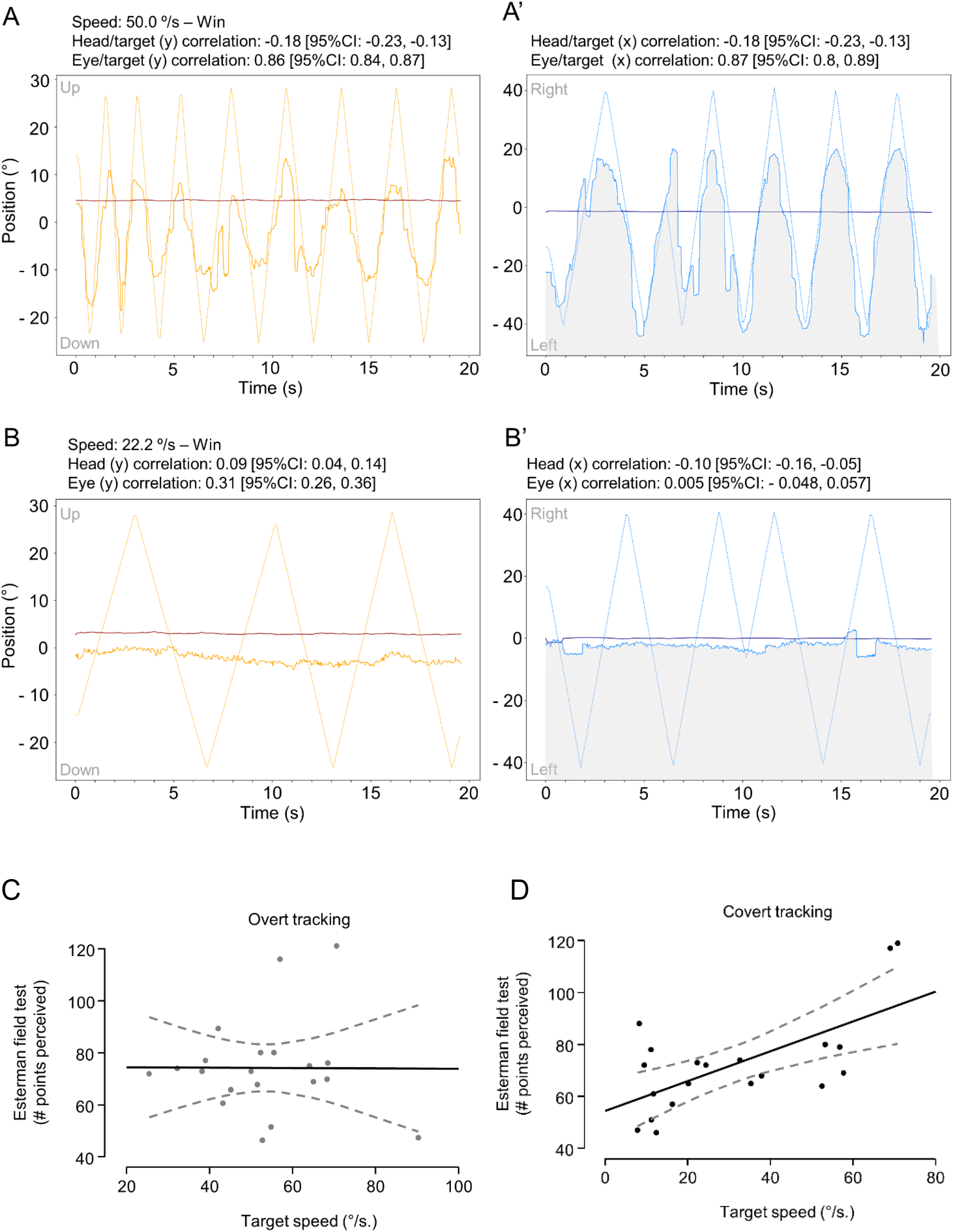
Correlation between tracking paradigms and number of points seen. Representative example of head/eye/target trajectory recordings during overt tracking in P9 (left homonymous hemianopia). **A- A’**. Graphs represent the position (in degree) as a function of time (s) of the target (orange (A) and blue (A’) dashed lines), the head (red (A), dark blue (A’) solid lines) and the eyes (orange (A) and blue (A’) solid lines) following the Y (orange, **A**) and X (blue, **A’**) axes during ***OVERT*** tracking. The grey shaded area represents the blind field. Head/target and Eye/target correlation coefficients for each axis are represented. **B-B’.** Graphs represent the position (in degree) as a function of time (s) of the target (orange (B) and blue (B’) dashed lines), the head (red (B), dark blue (B’) solid lines) and the eyes (orange (B) and blue (B’) solid lines) following the Y (orange, B) and X (blue, B**’**) axes during ***COVERT*** tracking. The grey shaded area represents the blind field. Head/target and Eye/target correlation coefficients for each axis are represented. **C, D.** Graphs represent the performance (speed of the target - in °/s - for a correct choice) as a function of the number of points seen at the Esterman field test during ***OVERT*** (**C**) tracking (target speed in °/s: 52.5 [25.4, 42.9, 64.3, 90.3], points perceived: 73.2 [46.0, 66.5, 76.8, 119]) and ***COVERT*** (**D**) tracking (target speed in °/s: 23.3 [7.1, 11.58, 48.9, 70.8], points perceived, 70.5 [46.0, 63.2, 78.2, 119]). Dashed lines represent the 95%CI.

We then explored the relationship between performance in the 3D-MOT-IVR and the Esterman binocular field test outcome. Across all 10 participants, the analysis of overt tracking performance (target speed for a correct choice) revealed no correlation with the points perceived in the Esterman binocular field test (Pearson’s r = −0.006, [95%CI, −0.45, 0.44], p = 0.981, Figure 4C). Conversely, during covert tracking, a robust correlation was observed between average performance and the points perceived in the Esterman binocular field test for all 10 participants (Pearson’s r = 0.65, [95%CI, 0.27, 0.85], p = 0.003, Figure 4D). Reading speed exhibited a weak correlation with overt tracking but not with covert tracking (r = 0.57, [95%CI, −0.09, 0.88], p = 0.08 and r = 0.12, [95%CI, −0.34, 0.53], p = 0.625 respectively).

## Discussion

This pioneering study investigated the feasibility, safety, and potential effectiveness of an innovative intervention, the 3D-MOT-IVR. This dynamic, personalized, home-based, and remotely controlled audiovisual stimulation targeted visual rehabilitation in young individuals with hemianopia resulting from a brain tumour. Our notable findings revealed that 50% of the participants (responders) improved the perception of static luminous stimuli in the blind field, as assessed by the Esterman binocular field test post-intervention, aligning with earlier research^13,14^. Meanwhile, the remaining 50% of participants were stable. This marks the first instance of improved visual perception in the blind field extending beyond central vision (> 15°), observed in children and adolescents with hemianopia due to brain tumours within an intervention, in the comfort of their home, lasting under 2 months and with benefits persisting for up to 6 months post-intervention.

Importantly, our results revealed a robust link between performance in the 3D-MOT-IVR covert tracking and the perceived points in the Esterman field test. In other words, better covert tracking led to more points perceived in the Esterman binocular field test, emphasizing the intervention’s effectiveness in restoring visual perception in the blind field. In addition, a better outcome in binocular field recovery was observed with a 6-week intervention compared to a 4-week intervention. Further analysis of the participants’ performance at the 3D-MOT-IVR stimulation correlated to gaze tracking revealed that they keep track of the target after it entered their blind field without moving their eyes or head. This suggests a probable reliance on the spatial sound emitted by the target, in line with earlier findings showing that covert tracking relies on vision and audition^36^. Both animal and human studies have indicated that restoring visual perception in the blind field after audiovisual stimulation relies on multisensory plasticity, reorganization and reinforcement of neuronal networks, and recalibration of spatial awareness between vision and audition in sub-cortical (superior colliculus and pulvinar controlling covert attention) and cortical areas (hMT+/V5)^37–42^. Our repeated audiovisual 3D-MOT-IVR stimulation, maintaining consistent background, occurrence rate, and spatial/temporal relationships between visual and auditory stimuli, maximizes neuronal reinforcement (following a Hebbian-like learning rule)^43^. This reinforcement leads to perceptual improvement, unlike real-world conditions where these features consistently vary in intensity and occurrence^43^.

The improvement at the Esterman field test after the intervention in 50% of the participants was not associated with the location of the tumour, the time from diagnosis nor an active treatment, leaving a gap in the identification of factors influencing the response to the intervention.

Feasibility and safety data showed that the audiovisual 3D-MOT-IVR can be performed over periods of 6 weeks, at home, as a routine integrated in real-world conditions and in a daily schedule of an adolescent’s life. An important parameter in perceptual learning approaches is the duration of the intervention which can extend up to 20 months^44^. Our results suggest that a minimum of 6 weeks of 3D-MOT-IVR stimulation is required to observe meaningful benefits on visual perception in the blind hemifield.

No adverse events were associated with the use of 3D-MOT-IVR, in line with previous work in children and young adults^31,45^. Our group of participants, all students, preferably performed the intervention between 15:00 and 21:00, presumably after school hours or summer camp, without affecting the performance at the 3D-MOT-IVR. The setup of the 3D-MOT-IVR stimulation was adjusted remotely through Wi-fi for each participant individually (personalized intervention) and all digital recording generated by the application were successfully sent through Wi-fi to our secured lab computers without loss of data. This indicates that data exchange through Wi-fi using remotely controlled Meta Quest2/Pro is reliable and safe, without data loss, further confirming that stand-alone, remotely controlled headsets are suitable devices for tele-/e-health in different patient populations^46^.

In contrast to previous reports on adults with hemianopia consecutive to stroke or brain trauma, contrast sensitivity in our participants was in the normal range before intervention but nonetheless clinically improved after intervention. Such discrepancy may be due to the type of brain injury (stroke, trauma versus tumour), the age of affected individuals (adults versus adolescents) and/or the method of assessment (FACT versus MRI)^47,48^. Reading speed showed clinically significant improvement after intervention as shown previously in our case series^13,14^.

This work, the first of its kind in a population of children and adolescents with hemianopia consecutive to brain tumour, demonstrated that a personalized, home-based, remotely controlled and dynamic audiovisual 3D-MOT-IVR stimulation lasting under 2 months can stably restore visual perception in the blind field, improving activities of daily living.

Case studies have shown that static, spatially and temporally correlated audiovisual stimuli displayed on computer screens can also improve perception in the blind field in adults/older adults with hemianopia after stroke, but require frequent clinic visits^49,50^. In addition, recent retrospective studies reported variable benefits of active Bevacizumab treatment on visual acuity and visual field in children with optic pathway glioma, although these improvements were often lost after discontinuation of the treatment^9–12^. The mechanisms underlying these temporary improvements are unknown. The effects of an intervention combining Bevacizumab and 3D-MOT-IVR should be explored. Our visual telerehabilitation program represents a paradigmatic shift in the conceptualization of healthcare, embracing a more holistic vision and facilitating the delivery of personalized care.

It ensures equitable access, offering the capability to receive supportive therapy at home independently, without direct supervision by healthcare professionals. This empowers children and adolescents residing outside urban areas to access specialized therapies. These advantages gain heightened significance in the recent context of the pandemic, given the challenges posed by travel restrictions, staff shortages, and related constraints, which have resulted in treatment delays for patients.

## Funding

Meagan Bebenek Foundation (E.B), University Health Network Foundation (M.R.)

## Conflict of Interest

The authors declare no conflict of interest.

## Authorship

Study design (E.B., M.R.), patient identification and recruitment (I.Y.B, A.R, U.T, U.B., V.R., E.B.), outcomes assessment (M.M, M.D.N., D.T., E.G.G, K.C., L.A., S.N.M.), clinical coordination (D.T., K.C.), data collection and analysis (M.M., B.Z., M.D.N, D.T., L.A., S.N.M, E.B., M.R.), application coding and virtual-reality implementation (E.G.G, P.N., K.C.), manuscript writing and editing (M.R, E.B., L.A., U.B., U.T.).

## Data Availability

Deidentified data and codes for data analysis are available upon request.

## Data Availability

All data produced in the present study are available upon reasonable request to the authors

## Acknowledgments

We thank the participants and their families for their participation and commitment.

## Notes

### Competing Interest Statement

The authors have declared no competing interest.

### Clinical Trial

NCT05065268

### Funding Statement

This study was funded by Meagan Bebenek Foundation and University Health Network Foundation.

### Author Declarations

The study protocol was approved by the Research Ethics Boards at the Hospital for Sick Children in Toronto, Canada (Sickkids REB# 1000076413) and at University Health Network, Toronto, Canada (UHN REB# 21-5978) and registered on clinicaltrials.gov (NCT05065268)

