## Supplementary files for "A telerehabilitation program to improve visual perception in children and adolescents with hemianopia consecutive to a brain tumour: a single-arm feasibility and proof-of-concept trial"

***Supplementary figures:***

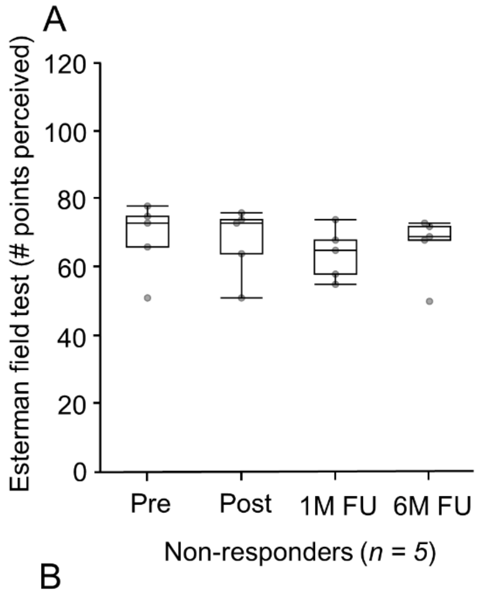


Pre: 73.0 [51.0, 66.0, 75.0, 78.0]
Post: 73.0 [76.0, 64.0, 74.0, 76.0]
1M FU: 65.0 [55.0, 58.0, 68.0, 74.0]
6M FU: 69.0 [50.0, 68.0, 72.0, 73.0]

| **Statistical analysis Esterman visual field test in non-responders (n = 5)** | | | | |
|---|---|---|---|---|
| | *Pre* | *post* | *post* | *1M* |
| | *post* | *1M* | *6M* | *6M* |
| *t* | (4)= 2.23 | (4)= -1.69 | (4)= -0.576 | (4)= 1.162 |
| *p* | 0.089 | 0.167 | 0.595 | 0.329 |
| $BF_{10}$ | 1.48 | 0.958 | 0.454 | 0.483 |
| $\delta$ | -0.7 | 0.52 | 0.17 | -0.21 |
| *[95%CI]* | [-1.82, 0.18] | [-0.28, 1.51] | [-0.54, 0.96] | [-1.02, 0.50] |

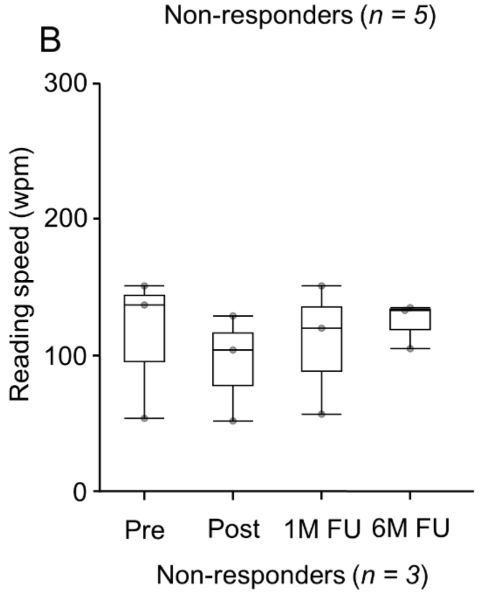


Pre: 138.0 [55.0, 6.5, 145.0, 152.0]
Post: 105.0 [53.0, 79.0, 117.5, 130.0]
1M FU: 121.0 [58.0, 89.5, 136.5, 152.0]
6M FU: 134.0 [106.0, 120.0, 135.0, 136.0]

| **Statistical analysis reading speed in non-responders (n = 3)** | | | | |
|---|---|---|---|---|
| | *Pre* | *post* | *post* | *1M* |
| | *post* | *1M* | *6M* | *6M* |
| *t* | (2)= -0.347 | (2)= 2.879 | (2)= -2.070 | (2)= 0.787 |
| *p* | 0.31 | 0.102 | 0.174 | 0.514 |
| $BF_{10}$ | 0.801 | 1.644 | 1.167 | 0.59 |
| $\delta$ | 0.459 | -0.934 | -0.688 | -0.273 |
| *[95%CI]* | [-0.47, 1.73] | [-2.97, 0.27] | [-2.30, 0.35] | [-1.34, 0.60] |

**Supplemental Figure 1**: **A**. Number of points seen and statistical analysis at the Esterman binocular field test in non-responders (n = 5/10) pre/post-intervention and at 1-month (1M FU) and 6-month (6M FU) follow-ups. **B**. Reading speed in words per minute (wpm) and statistical analysis in non-responders (n = 3/10) pre/post-intervention and at 1M FU and 6M FU.

A

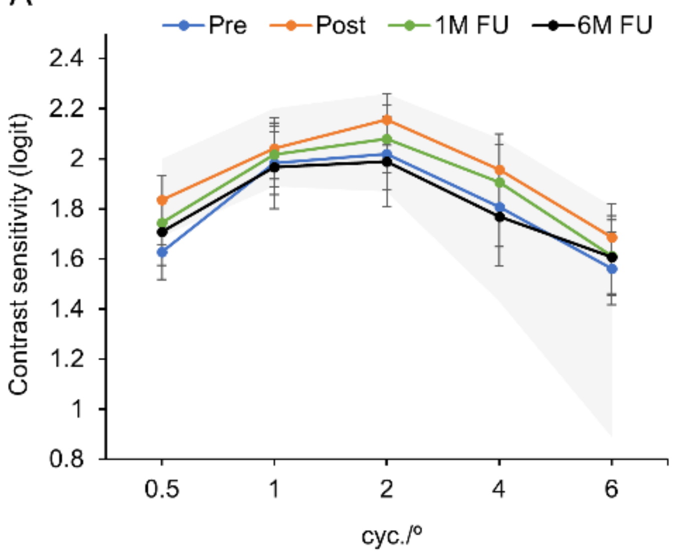


| Period | Logit difference for each cycle/degree | | | | |
|---|---|---|---|---|---|
| | ***0.5*** | ***1.5*** | ***2*** | ***4*** | ***6*** |
| **Pre-post** | 0.21 | 0.06 | 0.14 | 0.15 | 0.13 |
| **Post - 1M** | -0.09 | -0.03 | -0.08 | -0.05 | -0.07 |
| **Post - 6M** | -0.13 | -0.08 | -0.17 | -0.19 | -0.08 |
| **1M - 6M** | -0.04 | -0.05 | -0.09 | -0.14 | 0.00 |
| *COR ±* | *0.25* | *0.22* | *0.25* | *0.45* | *0.6* |

| Statistical analysis pre-/post-intervention (both eyes) | | | |
|---|---|---|---|
| *Cyc/°* | *$BF_{+0}$* | *δ* | *[95%CI]* |
| *0.5* | **3.4** | **0.46** | **[0.07, 0.92]** |
| *1* | 1.24 | 0.33 | [0.03, 0.77] |
| *2* | **6.4** | **0.53** | **[0.12, 1.02]** |
| *4* | **7.55** | **0.55** | **[0.13, 1.04]** |
| *6* | **4.91** | **0.50** | **[0.09, 0.98]** |

B

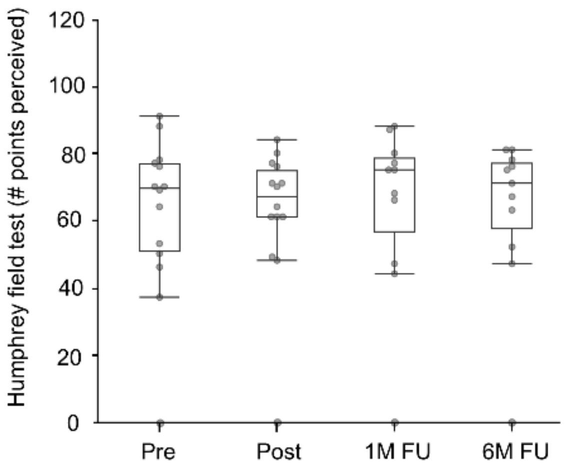


Pre: 69.5 [0.0, 50.7, 76.7, 91.0]
Post: 67.0 [0.0, 61.0, 74.7, 84.0]
1M FU: 75.0 [0.0, 56.5, 78.5, 88.0]
6M FU: 71.0 [0.0, 57.5, 77.0, 81.0]

| Statistical analysis Humphrey visual field test | | | | |
|---|---|---|---|---|
| | *Pre* | *post* | *post* | *1M* |
| | *post* | *1M* | *6M* | *6M* |
| *t* | (19)= 1.126 | (17)= -1.236 | (17)= -1.342 | (17)= 0.034 |
| *p* | 0.274 | 0.233 | 0.197 | 0.973 |
| *$BF_{10}$* | 0.405 | 0.469 | 0.525 | 0.243 |
| *δ* | 0.219 | 0.25 | 0.272 | 0.007 |
| *(95%CI)* | [-0.19, 0.64] | [-0.18, 0.70] | [-0..16, 0.73] | [-0.42, 1.43] |

**Supplemental Figure 2**: **A**. Contrast sensitivity measures and statistical analysis pre- (blue), post- (orange) intervention and at 1-month (1M FU, green) and 6-month (6M FU, black) follow-ups. Shaded area indicates the range of normal values. **B.** Number of points perceived and statistical analysis at the Humphrey field test pre/post-intervention and at 1- (1M FU) and 6-month (6M FU) follow-ups.

***Supplementary tables:***

| **Questions** | **Median (cutoff score: 5)** | **Min** | **q1** | **q3** | **Max** |
|---|---|---|---|---|---|
| 1. How was it to put the device on? | 6.5 | 4.0 | 5.25 | 7.0 | 7.0 |
| 2. How was the adjustment of the device using the straps? | 6.5 | 4.0 | 5.0 | 7.0 | 7.0 |
| *3. How was the adjustment of the device wearing glasses?* | *NA* | *NA* | *NA* | *NA* | *NA* |
| *4. How was the use of the device wearing glasses?* | *NA* | *NA* | *NA* | *NA* | *NA* |
| 5. How was the quality of the graphics? | 5.0 | 3.0 | 4.0 | 5.75 | 7.0 |
| 6. How was the quality of the sound? | 5.0 | 4.0 | 4.25 | 5.0 | 6.0 |
| 7. How was the quality of the VR technology overall (hardware, peripherals)? | 6.0 | 4.0 | 5.25 | 6.0 | 7.0 |
| 8. How was the use of the pointer? | 6.5 | 5.0 | 5.25 | 7.0 | 7.0 |
| 9. How was the selection using the pointer? | 5.0 | 4.0 | 4.25 | 7.0 | 7.0 |
| 10. How was the device's recharging process? | 7.0 | 5.0 | 6.0 | 7.0 | 7.0 |
| **Total score** | **47.5** (max: 56, cutoff: 40) | 33 | 39.25 | 51.75 | 55 |

**Supplemental Table 1:** Virtual Reality Neuroscience Questionnaire (VRNQ) – adapted.
Questions #3 and #4 were disregarded as non relevant. *NA*, non-applicable.

**A**

| Median best corrected visual acuity (BCVA) near (LogMAR) | | | | |
|---|---|---|---|---|
| | *Pre* | *Post* | *1M* | *6M* |
| *Median* | -0.097 | -0.097 | -0.097 | -0.097 |
| *Minimum* | -0.097 | -0.097 | -0.097 | -0.097 |
| *q1* | -0.097 | -0.097 | -0.097 | -0.097 |
| *q3* | 0.0 | 0.0 | -0.097 | -0.097 |
| *Maximum* | 2.4 | 2.4 | 2.4 | 2.4 |

| Statistical analysis BCVA near | | | | |
|---|---|---|---|---|
| | *Pre* | *post* | *post* | *1M* |
| | *post* | *1M* | *6M* | *6M* |
| *t* | (18)= -0.941 | (15)= 0.615 | (15)= 1.331 | (15)= -1.000 |
| *p* | 0.359 | 0.548 | 0.203 | 0.333 |
| $BF_{10}$ | 0.351 | 0.302 | 0.539 | 0.393 |
| $\delta$ | - 0.186 | 0.129 | - 0.282 | - 0.211 |
| *(95%CI)* | [-0.62, 0.23] | [-0.32, 0.59] | [-0.76, 0.18] | [-0.68, 0.24] |

**B**

| Median best corrected visual acuity (BCVA) far (LogMAR) | | | | |
|---|---|---|---|---|
| | *Pre* | *Post* | *1M* | *6M* |
| *Median* | 0.0 | 0.0 | 0.0 | 0.0 |
| *Minimum* | -0.097 | 0.0 | 0.0 | 0.0 |
| *q1* | 0.0 | 0.0 | 0.0 | 0.0 |
| *q3* | 0.376 | 0.279 | 0.301 | 0.301 |
| *Maximum* | 2.4 | 2.4 | 2.4 | 2.4 |

| Statistical analysis BCVA far | | | | |
|---|---|---|---|---|
| | *Pre* | *post* | *post* | *1M* |
| | *post* | *1M* | *6M* | *6M* |
| *t* | (18)= - 0.894 | (16)= 1.459 | (16)= 1.308 | (17)= 0.328 |
| *p* | 0.383 | 0.164 | 0.209 | 0.747 |
| $BF_{10}$ | 0.338 | 0.610 | 0.516 | 0.261 |
| $\delta$ | - 0.177 | 0.303 | 0.271 | 0.067 |
| *(95%CI)* | [-0.61, 0.24] | [-0.15, 0.78] | [-0.18, 0.74] | [-0.37, 0.51] |

**C**

| Fixation stability (63% BCEA) in $^{o2}$ | | | | |
|---|---|---|---|---|
| | *Pre* | *Post* | *1M* | *6M* |
| *Median* | 0.1 | 0.15 | 0.2 | 0.1 |
| *Minimum* | 0.1 | 0.04 | 0 | 0 |
| *q1* | 0.1 | 0.1 | 0.1 | 0.1 |
| *q3* | 0.325 | 0.3 | 0.275 | 0.4 |
| *Maximum* | 20.1 | 8 | 10.9 | 8.7 |

| Statistical analysis fixation stability | | | | |
|---|---|---|---|---|
| | *Pre* | *post* | *post* | *1M* |
| | *post* | *1M* | *6M* | *6M* |
| *t* | (19)= 1.280 | (17)= -1.404 | (17)= -1.348 | (17)= 1.203 |
| *p* | 0.216 | 0.178 | 0.195 | 0.246 |
| $BF_{10}$ | 0.474 | 0.562 | 0.529 | 0.454 |
| $\delta$ | 0.249 | - 0.28 | - 0.273 | 0.243 |
| *(95%CI)* | [-0.17, 0.68] | [-0.74, 0.15] | [-0.73, 0.16] | [-0.19, 0.69] |

**Supplemental Table 2**: **A, B.** Best corrected visual acuity (BCVA) values near (**A**) and far (**B**) pre/post-intervention, 1-month (1M) and 6-month (6M) follow-up and statistical analysis. **C**. Fixation stability pre/post-intervention, 1-month (1M) and 6-month (6M) follow-up and statistical analysis.
